# Knowledge, Attitude and Practices (KAP) of Antimicrobial Usage in in Kisii County, Kenya

**DOI:** 10.1101/2025.01.26.25321155

**Authors:** Emmanuel O Babafemi, Simion K Omasaki, Edward Mogoko, Philippa G McCabe

## Abstract

**Background:** Antimicrobial stewardship (AMS) policy is critical in tackling antimicrobial resistance (AMR). The information is available at the national level in Kenya about AMS policy but needed to be strengthened among the health workers and facilities at the county level. The aim of this study was to conduct a baseline assessment of the Knowledge, Attitude and Practices (KAP) on Antimicrobial Prescription and its Resistance Among Health Care Providers in Resource Poor Settings using Kisii County, Kenya as case study.

**Methods:** A paper-based survey was conducted among Public, Private Health facilities and Community Pharmacies in Kisii County between October and December 2023. The data collected from the respondents were put into Microsoft Excel spreadsheet for coding before being imported to RStudio for analysis.

**Results:** A total of 154 facilities across the 45 wards in Kisii County were included in the study. Most participants 111 (72%) agreed they have not received training and education on AMR and its implications on public health. Similarly,75 (49%) of the respondents said they have no formal procedures for all prescribers to conduct daily reviews of antibiotic selection until a definitive diagnosis and treatment duration are established. And that, they were not aware of AMS policy. However, they would be interested in workshops to educate them on AMR/AMS. Finally, on the level of awareness of the diagnosis and surveillance of AMR mechanisms, 248 (40%) responses said they were not or not very familiar with Methicillin-resistant *Staphylococcus aureus* (MRSA), Vancomycin-resistant Enterococci (VRE), Extended-Spectrum Beta-Lactamases (ESBLs), Carbapenemase-producing enterobacterales (CPE), Multidrug-resistant tuberculosis (MDR-TB) etc.

**Conclusion:** This study found limited level of AMS implementation/uptake across all the facilities studied in Kisii County, Kenya. The study identified challenges that require urgent action in strengthening AMS policy. AMS training and education would be more effective if they are designed to specific healthcare specialties and communities/facilities in Kisii County.

## Introduction

Antimicrobial resistance (AMR) is considered a global health and development challenge. It is rising the human and animal morbidity and mortality rates while increasing the prevalence of AMR organisms in the environment [1]. It is an emerging global challenge that is thought to account for more than 700,000 human deaths annually [2]. For decades, microbes, particularly bacteria, have become increasingly resistant to various antimicrobials. The World Health Organization estimates that AMR is responsible for a staggering number of deaths annually. A 2019 WHO report estimated that 1.27 million deaths were directly attributable to bacterial antibiotic resistance. In addition, WHO projections are even more alarming, that by 2050 AMR could cause 10 million deaths each year, incurring a cumulative economic cost of a staggering $100 trillion USD [3].

Estimate of bacterial AMR in WHO African region from clinical and surveillance data bases and national antimicrobial consumption data confirm a substantial burden for the current and future health of the continent [4]. African countries have inadequate laboratory facilities to monitor AMR which hugely contribute to increasing drug resistance globally. In fact, resistance has been reported for diseases such as typhoid, cholera, meningitis, dysentery, TB, malaria and AIDS. Between 2008 and 2011 cotrimoxazole resistance was reported against Vibrio cholerae from affected countries. In the contrary, same pathogens remained extremely sensitive to quinolones and cyclines [5].

The burden of antimicrobial resistance (AMR) in Kenya goes further to confirm AMR as a global challenge. In Kenya, there’s no readily available data on the exact number of deaths attributable solely to AMR, but studies suggest a concerning situation. The 2019 WHO report on global AMR estimates that 8,500 deaths in Kenya were attributable to AMR, with an additional 37,300 deaths associated with AMR [6]. Kenya has the 177th highest age-standardized mortality rate per 100,000 populations associated with AMR across 204 countries. These numbers highlight the potential impact of AMR on mortality in the country. Consequences of irrational treatment and erroneous diagnosis, unregulated antimicrobial use, knowledge gap in antimicrobial courses, side effects and dosage limits are found to be the leading causes of AMR [7]. AMR has resulted to hard-to-treat infections with previous working antibiotics depicting unforeseen future in health care [8]. In addition, AMR has impacted serious infections and long hospitalizations, increased treatment failures and increased costs in healthcare [9]. Irrational use of antibiotics is catalysed by buying over the counter antibiotics without prescriptions. LMIC private retail chemists are potential primary patient care areas for self-prescribed antibiotics among patients [10]. Inappropriate antimicrobial use in LMICs hospitals has been fuelled by poor antimicrobial stewardship activities, deficit surveillance and diagnostic facilities and lack of antimicrobial treatment guidelines [11]. In LMICs, the AMR is worsened by: (i) inaccessible to rational antimicrobial therapy [12]; (ii) feeble regulations in the treatment and dispensing of antimicrobials for humans [13]; (iii) feeble audits and surveillance of AMU and AMR levels [13]; (iv) standard treatment guidelines that are not updated [14]. Inadequate community knowledge, poverty, lack of healthcare services has potentiated to inappropriate antimicrobial use (AMU) [15]. With the rates of AMR increasing worldwide and very few new antibiotics being developed, existing antibiotics are becoming a limited resource. It is therefore essential that antibiotics only be prescribed and that last-resort antibiotics be reserved for patients who truly need them. This responsible use is the foundation of the concept of antimicrobial stewardship (AMS).

According to the World Health Organization (WHO), antimicrobial stewardship is defined as “A battery set of actions which fuels the responsible use of antimicrobials. This definition can be actionable at the personal level as well as national and global level, and across human, animal health and the environment” [16]. This was originally coined for application in the hospital setting to optimise antimicrobial use, leading to the term "antimicrobial stewardship" (AMS) [17]. The idea has since enlarged to incorporate the governance of the health sector as a whole, accenting responsibility for population health and comfort, managing health systems at national and global levels [17]. Today, the three pillars of an integrated approach strengthening health systems, are AMS, infection prevention and control (IPC) and medicine and patient safety. Linkages of these pillars to other important activities of infection management and robust AMR surveillance and a dependable quality medicines supply, will ultimately contribute to the goal of achieving universal health coverage [18]. The World Health Organization (WHO) announced AMR as a major global problem and started a global action plan to combat the challenge [2]. Therefore, enhancing knowledge and surveillance skills was one of the parts of this action plan endorsing a universal antimicrobial resistance surveillance system. The WHO’s global strategy highlighted the need for appropriate antibiotic use along with the critical role of healthcare workers [19]. Effective implementation of the plan will help control AMR by promoting the careful use of antimicrobials [18].

Health care providers including doctors, pharmacists, and nurses play a key role in tackling antimicrobial resistance because they have the insight and knowledge to prescribe and dispense antibiotics during clinical practice as well as in promoting patients’ compliance to therapies and avoid self-medication [20]. Unfortunately, the misuse and overuse of antibiotic prescriptions by health care providers has been found to be a problem [21]. Roughly, studies have reported that 30-50% of patients in hospitals have incorrect indication for antibiotic, the antibiotic of choice or the dosage limit. Study in China showed that 30-60% of intensive care units cases, had inappropriately prescribed antibiotics [22]. Minimizing the irrational use of antibiotics is the best way to curb the spread of AMR, and this can be done by behaviour change in prescribing and dispensing practices among healthcare providers.

Evidence focused questionnaires like knowledge, attitude and practices (KAP) surveys can be used to analyse the factors influencing health care providers’ prescribing and dispensing behaviours [23]. In recent past, studies have shown insufficient knowledge on rational use of antibiotics among Chinese medical undergraduate students [24]. In addition, the medical students would self-prescribe unnecessary antimicrobials for infections [25]. Furthermore, a study in United States of America showed that knowledge and attitude regarding antibiotic use differed among antibiotic Hispanic consumers and non-Hispanic consumers. In fact, Hispanic consumers believed antibiotics could cure colds, thereby self-prescribing antibiotics, as well as obtaining antibiotics from grocery stores, family and friend or using earlier illness unconsumed antibiotics [26]. Self-prescribing is associated with increased antibiotic resistance as you miss appropriate judious provider care and opt to visit grocery shops to buy over-the counter antibiotics [27]. In India, one study reported that laypeople had rough idea and knowledge about AMR and AMS [28]. In contrary, a study done in Egypt showed health care providers had a good knowledge and attitude about AMS [29]. In addition to that Nigerian study conducted in tertiary hospitals reported that knowledge and attitude of antimicrobial prescriptions and AMR is wanting worsened by physicians’ inappropriate antibiotics prescriptions [30].

Furthermore, one particular study conducted in Bhutan strongly supported the need to involve the community pharmacies as participants in training and policy making processes to help address growing threat of antimicrobial resistance [31].

Therefore, we conducted a KAP survey to evaluate the awareness of antimicrobial usage and antimicrobial resistance among public and private tertiary care hospitals (levels 3,4 & 5) in Kisii County, Kenya, as such important information is lacking in Kisii.

This KAP survey on AMS, is aimed at addressing the growing threat of AMR in the region through understanding the health care providers’ knowledge, attitudes and practices on prescription and dispensing of Antimicrobials. This data will go a long way in enhancing insightful knowledge and understanding of AMR among healthcare workers in Kisii County through effective communication, education, and training programs.

Nevertheless, strengthening the existing knowledge and evidence-based practices in our region will be through conducting consistent surveys, surveillance and research activities within the Kisii County healthcare system and promoting the optimisation of antimicrobial use to public health sector.

## Methods

### Study Area

Kisii County is one of the 47 counties in Kenya courtesy of the new constitution of Kenya enacted in 2010, which created the new county system of governance. It shares common borders with Nyamira County to the North East, Narok County to the South and Homabay and Migori Counties to the West.

The county lies between latitude 030° and 10° South and longitude 34.38° and 35.0° East. The county covers a total area of 1,332.7 Km square and is divided into nine sub counties: namely Kitutu Chache South, Kitutu Chache North, Nyaribari Masaba, Nyaribari Chache, Bonchari, Bobasi, Bomachoge Chache, Bomachoge Borabu and South Mugirango. The county has 45 wards, which are the lowest units of administration.

According to the 2019 Kenya Population and Housing Census, the county had an estimated total population of 1,332,174 in 2022 comprising of 661,680 males and 670,494 females with a population density of 957 persons per square kilometre. The population is served by 32 community health units (level 1), 84 dispensaries (level 2), 28 health centres (level 3), 14 sub county hospitals (level 4) and 1 referral hospital –Kisii Teaching and Referral Hospital (level 5). The population is also served health facilities that are managed by private practitioners and faith-based organizations.

### Study Design and Site

This was a descriptive cross-sectional study that was conducted in Public and Private Health facilities and Community Pharmacies in Kisii County between October and December 2023. It utilised a quantitative data collection method using a paper-based survey that was applied through a purposive and random selection process of study areas and participants.

### Ethics statement

The study received ethics approval from the Institutional Scientific Ethics Review Committee (ISERC) of the University of Eastern Africa Baraton (Application approval number: UEAB/ISERC/02/10/2023). The informed verbal consent was obtained from all study participants. The responses and identities of key informants and survey participants were kept strictly confidential.

### Sample size

The sample size was drawn through a mixed sampling procedure as detailed below:

#### Community Pharmacies, Public and Private Health Facilities

The community pharmacists were chosen using simple random sampling method from 225 registered community pharmacies in Kisii County at the time of the study as obtained from the Pharmacy and Poisons Board register. From the 225 registered pharmaceutical outlets, a random sample of 40 pharmacies were picked to participate in the survey. For the health facilities, 4 facilities were purposively sampled from each of the 9 sub counties giving a total of 36. Criteria for their choice were that they must be levels 3, 4 or 5 (Health Centre, Sub County of County Referral Hospitals) as this is where we expected antibiotics is usually dispensed. The respondents were mainly the patients visiting the facility and health workers including doctors, nurses, clinical officers, laboratory technologists etc.

### Data collection

Before the commencement of data collection exercise, eighteen research assistants, mainly pharmacists or pharmaceutical technologists were trained on the study objectives, study protocol, sampling strategy and ethical issues. They were provided with visual aids including pictures of antimicrobials to make interactions with study participants clear or easy to understand. Data collection was done using a structured self-administered questionnaire. The questionnaire was pre-validated for its completeness, simplicity, accuracy, clarity, understandability, and relevance by experts in the field of clinical medicine and pharmacy. This gave room for modification of the initial questionnaire. The data collected included socio-demographic characteristics (age, sex, occupation, village, workplace, and residence), work experience, knowledge of antimicrobials (what are they and their roles, types of antimicrobials known and health conditions treatable with antimicrobials), practices of AMU (preferences, frequency of use, sources, ill-health conditions treated, adherence to the treatment regimen, self-medication practices). We also assessed awareness and knowledge of AMR. In addition to dichotomous responses to some questions, the five-point Likert scales (‘strongly agree’, ‘agree’, ‘uncertain’, ‘disagree’ and ‘strongly disagree’) were used to determine the participants’ knowledge and attitudes regarding antimicrobials, their uses as well as the burden, actions and roles to address AMR. Dependent variables included knowledge, attitude, and practices. Independent variables included age, sex, level of education, and work experience.

### Data Management and Analysis

The data collected from the respondents were put into MS Excel spreadsheet for coding before being imported to RStudio for data cleaning, statistical analysis and generation of graphs. Some graphs were also created in MS Excel spreadsheet. The descriptive statistics and graphs were used to demonstrate the current landscape regarding antibiotic usage within the regions and facilities investigated.

Responses to the triad of knowledge, attitudes and practices (KAP domains) were assessed using a scoring scheme. For dichotomous responses, a right response was given one score, and zero to the wrong/unanswered option. Multiple responses to each of the correct options were given one score; otherwise, no score was given. Responses to the five-point Likert scale were given scores that ranged from “5” for the most appropriate answer to “1” for the least appropriate answer and summed to form a discrete variable.

## Results

### Demographics

This study was conducted using three different surveys across Kisii County, Kenya, Africa to investigate the use of antibiotics and determine if an antimicrobial stewardship programme would be of use and value.

**Table 1:**
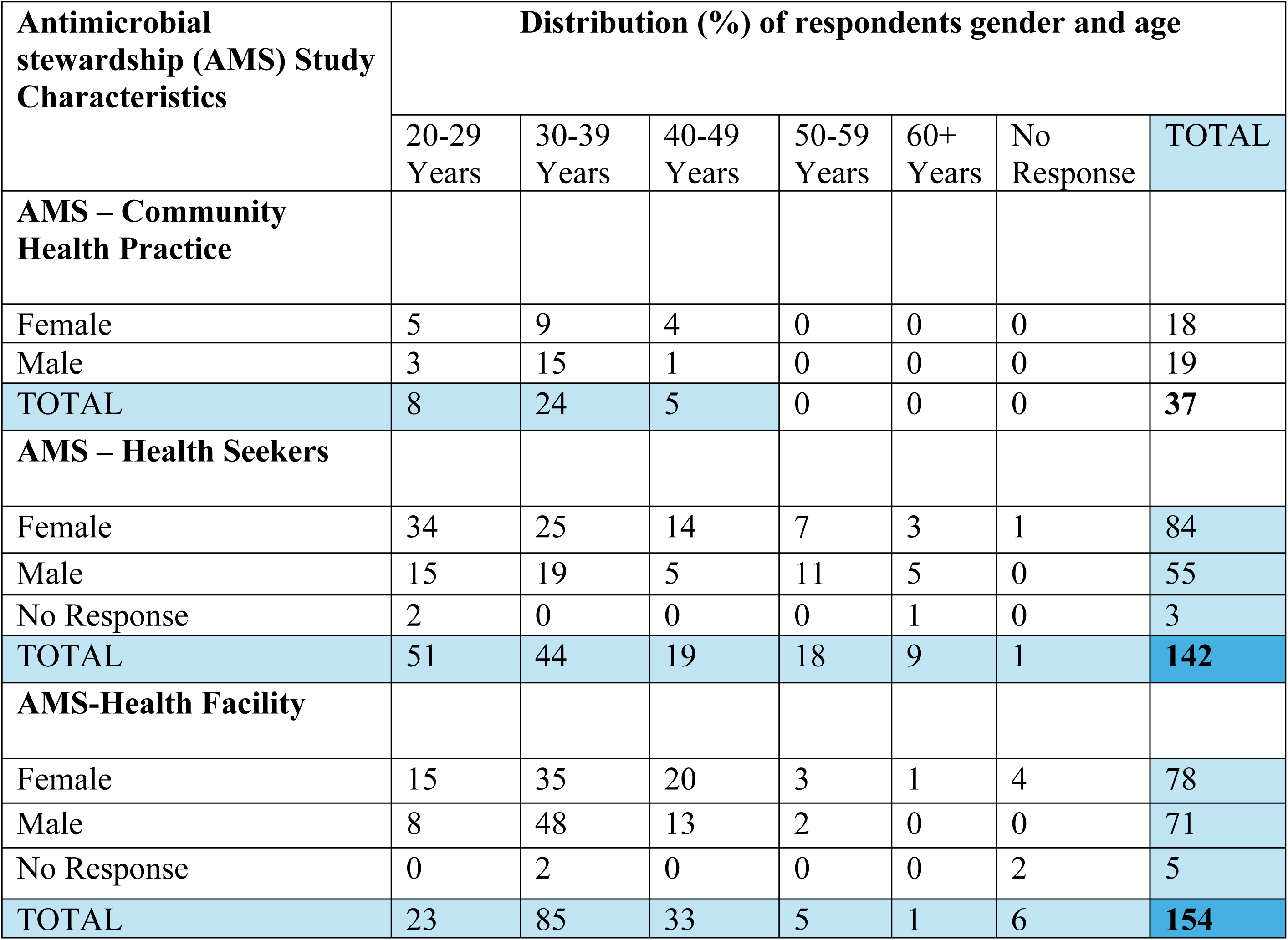
Showing the ages and sex distribution of the respondents to the survey.

### Exploratory Data Analysis

To understand the data fully some data explorations were conducted. The purpose of this was to highlight and identify any patterns in the data. Figure 1 shows the most surveyed age bracket is the 30 – 40-year age group.

**Figure 1.**
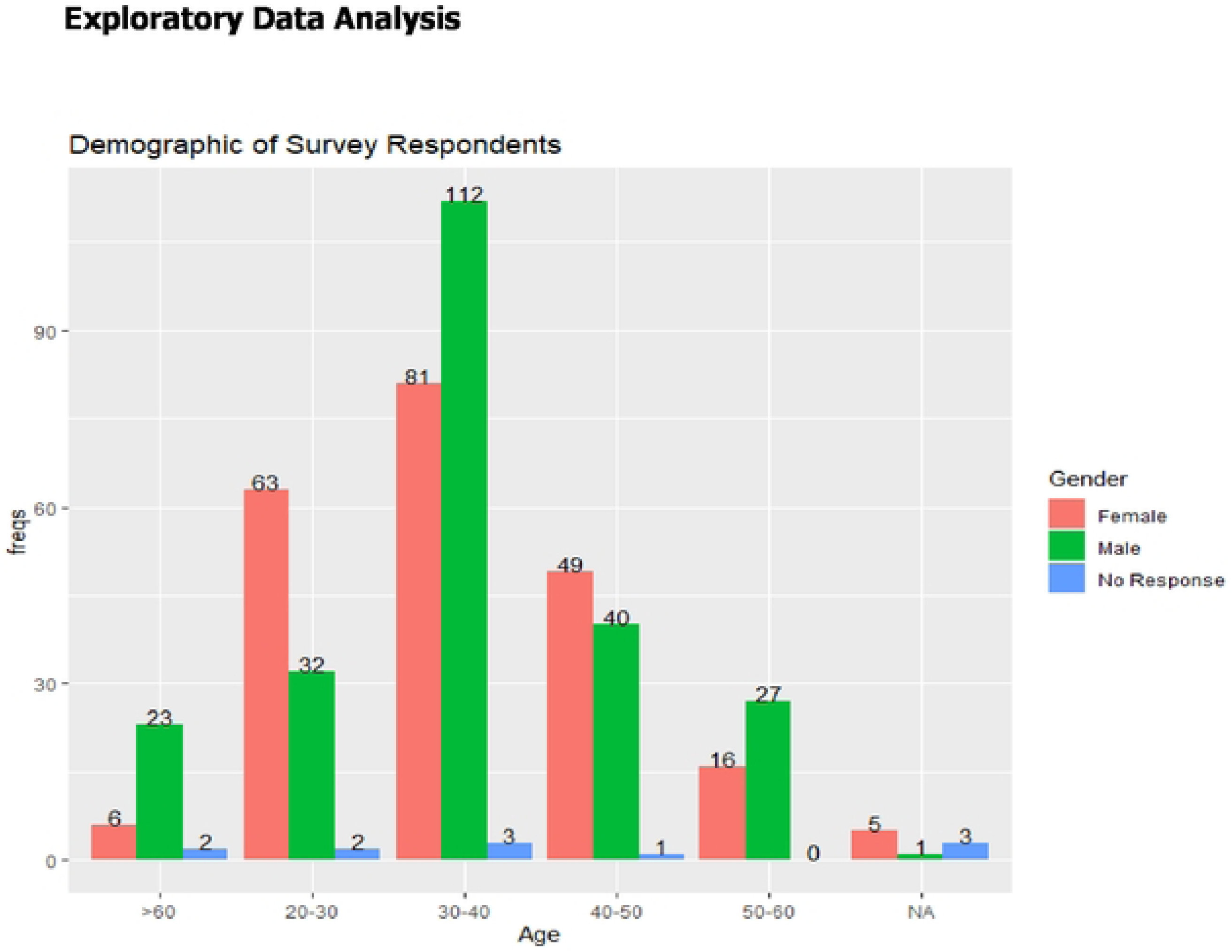
Demographic survey of the respondents.

### Quantitative findings

A total of 154 facilities across the 45 wards in Kisii County were included in the study. These are Faith-based, 21 (14%); Public, 26 (17%); Private, 95 (61%); and other facilities, 12 (8%).

### Community Health Practice

Data were collected across 16 wards, and 20 sub counties. The respondents of the survey reported on their professions. The responses to the survey show that 3 (8%) of the pharmacies are in major towns, 11 (30%) are in market centres and 21 (57%) are in small towns. There were 2 (5%) respondents who did not report on the location of their pharmacies. The pharmacy ownership was recorded in the survey at 21 (57%) respondents owning their pharmacy and 16 (43%) respondents not owning a pharmacy. When asked if they had received training, 19 (51%) said they had not, with 14 (38%) said they had, and 4 (11%) did not respond.

### Health Seekers

Data were collected across 42 wards, and 26 sub counties. Of the respondents, 4 (11%) people did not respond to the question about maximum education, 30 (81%) people reported diploma level, and 3 (8%) said that bachelor’s level was the highest education they had received. Furthermore, the respondents from this category included health seekers and health providers across Level 3, 32 (21%); Level 4, 15 (10%) and Level 5, 105 (68%) whilst 2 (1%) respondents did not indicate their levels.

### Health Facility

Data were collected across 40 wards, and 28 sub counties. The number of these facilities were 21 (14%), 26 (17%), 95 (61%) and 12 (8%) for Faith-based, Public, Private and others respectively. The respondents across the facilities were 32 (21%) from Level 3; 105 (68%) from Level 4; 15 (10%) from Level 5 while 2 (1%) respondents did not indicate their level. The distribution of positions of healthcare professionals were Clinical officers 47 (30%), Doctors 3 (2%), Nurses 47 (30%), Pharmaceutical technologists 21 (14%), Pharmacists 4 (3%) and others 32 (21%). These healthcare workers’ years of experience are less than 3 years-35 (23%), between 4 and 9 years-64 (42%); 10-14 years-36 (23%); 15-19 years-8 (5%) and greater than 20 years-11 (7%).

### Medical insurance

Of the 142 survey responses, 83 (58%) said they have National Health Insurance Fund (NHIF) Cover, 58 (41%) said they did not and 1 did not respond. When asked if they have private medical insurance, 2(1%) respondents did not answer, 7 (5%) said yes, but 133 (94%) said they did not.

### Self-medication

Of the survey respondents, 60 (42%) said they do treat themselves if they are unwell, 81 (57%) said they do not and 1 (1%) did not respond. Of those who have treated themselves, 26 (43%) said they have done this after a test, and 34 (57%) said they treated themselves before a test. Of the 60 responses that said they self-treat illness, 3 (5%) said they always do it, 18 (30%) said they often do it, 36 (60%) said they do it occasionally and 3 (5%) said they were unable to remember the frequency.

Of those people surveyed, 92 (65%) said they know about antibiotics, 44 (31%) did not and 6 (4%) gave no response. When asked if the person self-medicating with antibiotics checked the medication instruction leaflet 95 (70%) said they always check, 34 (24%) said they sometimes check, 8 (6%) said they were not sure, 1 (1%) was not given instructions and 4 (3%) did not respond.

When asked if they were aware of the regulations surrounding self-medicating when using antibiotics 63(44%) said yes, they were aware, 76 (54%) said they were not aware and 3 (2%) did not respond.

When asked if they knew, what the correct dosages were, 3 (2%) did not respond, 102 (72%) said yes, they were aware and 37 (26%) said they were unaware of the correct dosage.

The respondents of the survey reported that 92 (65%) knew the appropriate duration to take antibiotic medication, 45 (32%) do not know the right duration to take antibiotics for, and 5 (3%) did not respond.

When asked if they would recommend the treatment, 41 (29%) said they would, 87 (61%) said they would not and 14 (10%) did not give a response.

When asked if they would normally use similar drugs when unwell, 39 (27%) said they would, 96 (68%) said they would not and 7 (5%) gave no response.

### Awareness of Antimicrobial Resistance (AMR) Mechanisms

When asked if the individual has ever encountered patients infected with MRSA, VRE, CPE and MDRTB. The responses from the survey reported that 332 (54%) were somewhat or very familiar, 248 (40%) were not or not very familiar with MRSA, VRE, CPE MDRTB etc while with 36 (6%) no responses were recorded.

### Awareness of Antimicrobial Stewardship (AMS) Policies

Out of the 154 respondents on the level of awareness of the AMS; 85 (55%) people said no, 57 (37%) people said yes, and 12 (8%) people did not respond.

When asked how important is it for healthcare professionals to be aware of the diagnostic methods for detecting AMR, 101 (66%) said it was extremely important; 42 (27%) said it was very important; and 7 (5%) gave no answer at all whilst 2 (1%) said they didn’t know, and 2 (1%) said that it was not important at all to be aware of methods used to detect AMR.

When asked does your facility have policy that requires prescribers to document in the medical record or during order entry a dose, 9 (6%) did not respond, 31 (20%) said no and the remaining 114 (74%) said yes.

### Test-before-treat Protocol

When asked if they were aware of guidelines or policies related to testing before treating for infectious diseases 12 (8%) did not offer any response, 38 (25%) said they were unaware whilst the remaining 104 (68%) said that they were aware of guidelines.

When the respondents were further asked whether they feel that the guidelines are effectively implemented 18 (12%) said they were not, 79 (51%) said they were partially implemented, 23 (15%) said they were fully implemented and 34 (22%) did not respond.

Another question was asked if a ‘test-before-treat’ system could be used in practice to reduce AMR, 3 (2%) said no it would not work, 3 (2%) said they were unsure if it would work, 140 (91%) said yes, they thought this would help and 8 (5%) provided no response.

### Presence of Formal Prescribers Procedures

When asked if the facility have formal procedures for all prescribers to conduct daily reviews of antibiotic selection until a definitive diagnosis and treatment duration are established, 67 (44%) responded yes there are formal procedures, 75 (49%) said there were no such procedures, 10 (6%) did not respond and 2 (1%) said this was not applicable.

**Table 2:**
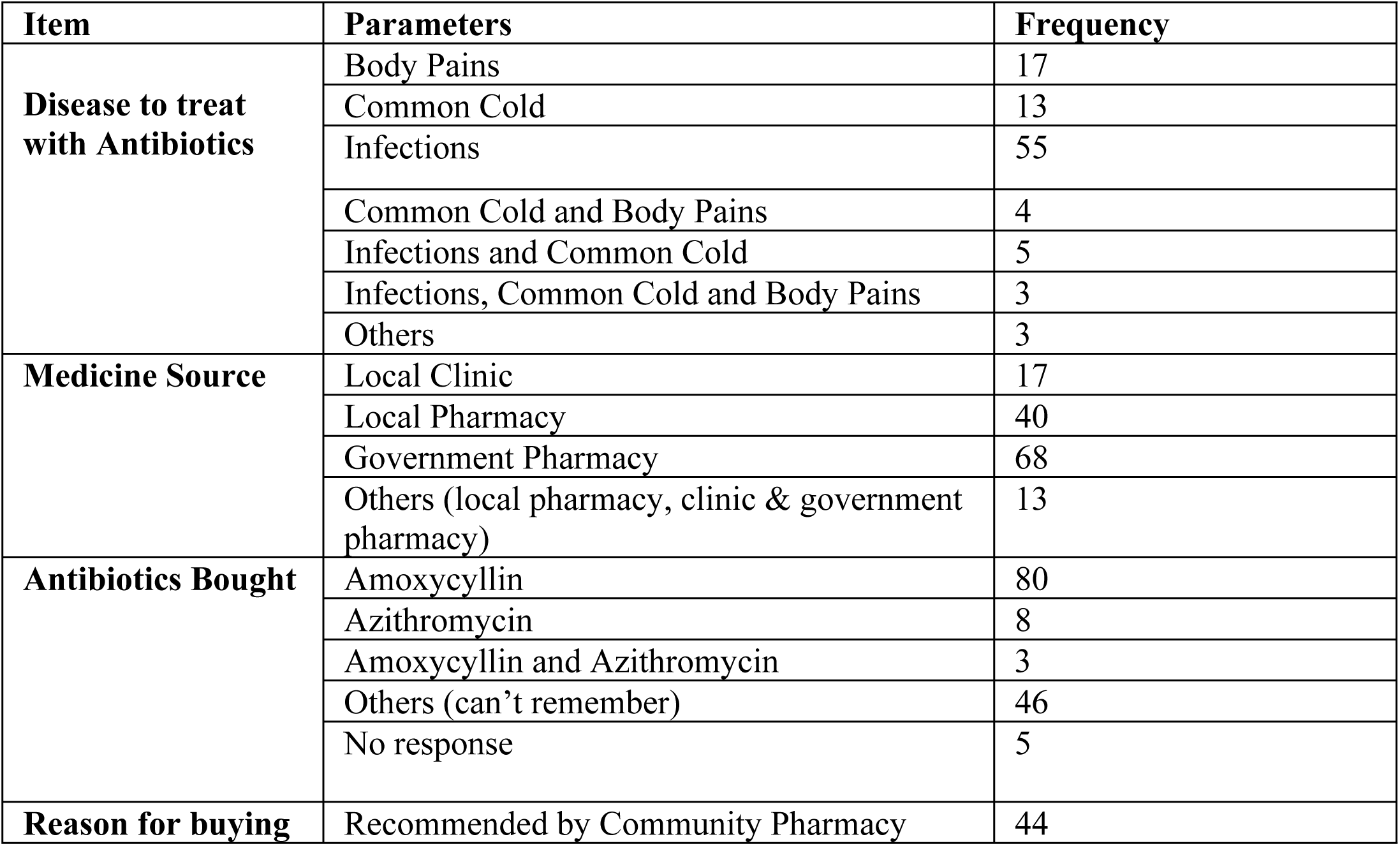
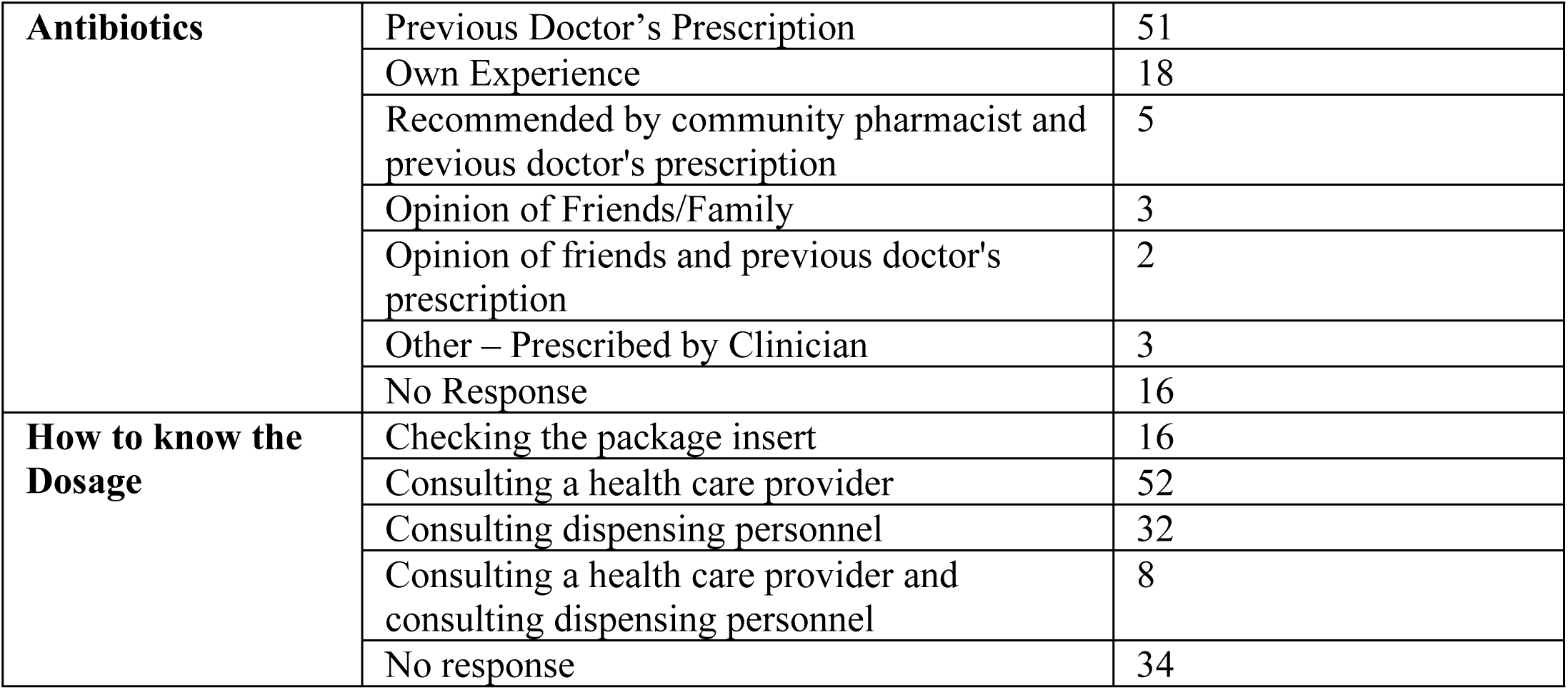
Showing Antibiotics usage among the respondents.

### AMR Training/Education

When asked if they have received any training/education on antimicrobial resistance and its implications on public health, 111 (72%) said no they had not received any training, 33 (21%) said they had received training/education, whilst the remaining 10 (7%) gave no response.

As a follow-on question, asking if the subjects would be interested in participating in workshops 8 (5%) gave no response, 5 (3%) said no, 3 (2%) said maybe and the remaining 138 (90%) said they would be interested in workshops.

## DISCUSSION

This study assessed the Knowledge, Attitude and Practices (KAP) on Antimicrobial Prescription and its Resistance Among Health Care Providers and workers in Kisii County, Kenya. This study indicated that none of the assessed hospitals and other health facilities had fully functional AMS policy.

Another major finding from our study was that most respondents admitted their antibiotic usage without prescription for the treatment without awareness of regulations surrounding self-medication and no evidence for infection. The troubling report was about a third (29%) of the respondents said they would recommend their treatment to others. Other findings from our study are respondents using similar drugs when unwell without knowing the right duration to take the antibiotics. The findings from the study further highlighted the serious problems around the antibiotic abuse/misuse. Antibiotics are readily available as over the counter medication that can be accessed without prescription. All these highlight the challenges confronting AMR/AMS in some LMICs settings [32, 33, 34]. While 54% of the responses agreed that they are somewhat or very familiar with importance of AMR mechanisms; 40% of the responses were not or not very familiar with MRSA, VRE, ESBL, CPE, MDRTB etc. while 6% did not respond. These findings reflected the widely endorsed, deeply embedded systemic infrastructural deficiencies in the diagnosis and microbiological surveillance of guiding the effectiveness of AMS in those settings [35, 36]

This finding highlights the need for the provision of resources that will strengthen the uptake of AMS and encourage its implementation across the different levels of the health facilities in Kisii county. Lack of resources have been identified as a risk to the successful implementation of AMR and AMS programs [37]. Another finding identified was the issue associated with a lack of leadership commitment in Kisii county, Kenya which has prevented the establishment of robust AMS programmes in hospitals and other health-based facilities [38]. Other studies have reported the importance of leadership commitment for a successful implementation of AMS policy. The absence generally affects implementation, accountability, responsibility, and commitment to AMS programmes [39, 40, 41]. Therefore, effective leadership is essential from Kisii County Government and the facilities management that would facilitate the implementation of issues around quality, diagnosis, surveillance, provision of infrastructures and other sustainable components of AMS activities [39]. It is therefore relevant for the leadership commitment and accountability as essential components in establishing and implementing effective AMS policy in hospitals and other health facilities [39, 40].

Lack of implementation of the AWaRe tool was noted and this may lead to inappropriate prescribing and use of antibiotics [42, 43]. Evidence has indicated that the lack of implementation of the AWaRe tool in hospitals and other facilities in the study could be due to inadequate awareness among healthcare workers and providers [44]. Therefore, increasing awareness, training and education about the AWaRe framework of rational antibiotics prescription and usage can improve adherence levels [44, 45]. It was observed that, the lack of implementation of AMS policy in hospitals and other health facilities could affect the rational use of antibiotics [46].

Therefore, there is a need to establish and strengthen AMS policy uptake in the hospitals and other health facilities in Kisii County. This will optimise and address the challenges of indiscriminate antibiotic use, improve patient outcomes and tackle the menace of AMR [47, 48, 49, 50].

Our study revealed that most facilities had challenges in conducting tests before treatment across the healthcare facilities. However, from the findings of our study and the assurances of the Kisii County Government, the implementation of AMS policy and activities will be encouraged across private, public and faith-based health facilities in Kisii county. Therefore, the identified gaps can be used as catalyst to strengthen AMS policy and activities in subcounty, county and referral tertiary hospitals in Kisii county to optimize the appropriate use of antibiotics in hospitals and the community.

The study strongly recommends to the policy makers for the establishment of AMR detections, strengthening of the existing AMS policy and National Action Plan (NAP) through active implementation across the hospitals and all levels of healthcare provisions in Kisii county.

One of the strengths of this study was engaging local stakeholders, including senior management, frontline healthcare professionals, and public health specialists, thus increasing elements of co-creation and a subsequent sense of ownership for the materials. Furthermore, our selection strongly considered representativeness as it engaged stakeholders across all partnership sites in all sub counties in Kisii. Additionally, we incorporate open-ended questions within the AMS checklist to allow for more reporting flexibility.

The study site (Kisii county) could be considered a limitation considering 47 counties in Kenya; however, results are transferable and that the approach can be implemented within other counties in Kenya and other Sub-Saharan Africa where implementation of AMS policy needs strengthening or non-existent.

Another limitation is the fact that, the pre-validated questionnaires consist of different questions, it is not possible to compare each of the cohorts to the others for the statistical analysis, however demographic information collected could be compared, allowing for a picture of the studied population to be considered (Figure 1).

Finally, as the study was conducted in Kisii county, however, some of the findings might not be generalisable to other counties in Kenya with better implementation of AMS policy in place. However, the protocol and methodology we used can be adapted in other counties to identify systematic obstacles and strengthening the specific AMS policy targets for LMICs

### Conclusion

This study revealed critical gaps in AMS policy in Kisii County Government Health System as demonstrated across the selected facilities. There was lack of resources, education and training on AMR and AMS, low level of leadership commitment to AMR diagnostic testing and AMS activities within healthcare facilities, poor documentation of AMS policy in Kisii County. We recommend an urgent need for the Kisii County Government to budget sufficient funding for interventions for AMS programmes across all healthcare facilities in the county.

There is an urgent need to mobilise domestic funding, design targeted interventions for AMS programs in healthcare facilities, and build capacity among healthcare workers and hospital leadership regarding all AMS core elements.

## Data Availability

All relevant data are within the manuscript and its Supporting Information files.

## Authors Contributions

EB (Emmanuel Babafemi): Conceptualisation, Methodology validation, Writing—original draft Writing and Review Project management/administration,

SI (Simon Omasaki): Data collection, cleaning and tabulation

EM (Edward Mogoko): Data collection, Writing-editing and Review PM (Philippa McCabe): Statistical analysis

All authors have read and agreed to the published version of the manuscript.

